# Google Trends as a method to predict new COVID-19 cases

**DOI:** 10.1101/2021.03.12.21253452

**Authors:** Tado Jurić

## Abstract

In this paper, we develop a method that can detect and predict the emergence of new cases of COVID-19 at an early stage. With this method, we try to lay the empirical basis for the development of the model of digital monitoring and prediction of the occurrence of new cases of COVID-19 in Croatia, relying on the analytical tool Google Trends (GT).

**Results:** In Croatia search activities using GT for terms such as ‘‘PCR +Covid”, “PCR + test”, and symptoms “cough + corona”, “pneumonia + corona”; “muscle pain + corona” correlate strongly with officially reported cases of the disease. Google Trends tools are suitable for predicting the emergence of new COVID-19 cases in Croatia, and that the data collected by this method correlate with official data.

The benefit of this method is reliable estimates that can enable public health officials to prepare and better respond to the possible return of a pandemic in certain parts of the country. If a region experiences an early, sharp increase in Covid-19-like-illness Google searches, it may be possible to focus additional resources on that region to identify the etiology of the outbreak, providing extra medical capacity or raising local media awareness as necessary.

Because the relative frequency of certain queries is highly correlated with the percentage of physician visits in which a patient presents with Covid-19 symptoms, this method can serve as an early alarm to predict the emergence of new cases of COVID-19 in the specific area in Croatia.

## Introduction

The Covid-19 outbreak and lockdown accelerated the adoption of digital solutions at an unprecedented pace, creating unforeseen opportunities for scaling up alternative approaches to social science. But it also brought digital risks and threats that placed new demands on policymakers (Hantrais et al, 2020).

Schwab and Malleret (2020) argued that, that the world today is “facing a ‘defining moment’ as the pandemic precipitated the fusion of technologies, enabling digital technologies to extend their reach, almost uncontrolled, into every aspect of life. Building on the third digital revolution, the Fourth Industrial Revolution is distinguished from previous industrial revolutions by its ‘velocity, scope, and systems impact’. This Fourth Industrial Revolution would develop exponentially rather than linearly and would ‘fundamentally alter the way we live, work, and relate to one another.” (Schwab, 2015). At the micro-level, families are shown to have become “digital by default” (Hantrais et al, 2020).

By mid-2020, 58% of the world population was estimated to be internet users, compared to almost 90% in the European Union (Internet World Stats, 2020). Within the EU, the same study showed that usage ranged from nearly 98% in Denmark to less than 70% in Bulgaria - in Croatia (79%) (Europa.eu, 2020).

The pandemic accelerated the uptake of digital solutions in data collection techniques (Hantrais et al, 2020). Budd et al. (2020) portrayed digital technologies being harnessed to support public health responses to Covid-19 worldwide. In the short term, face-to-face survey interviews were replaced by online interviewing, and by turning to other data sources. This direction of travel had to be abruptly scaled up as it became the “new normal” for data collection and dissemination (Hantrais et al, 2020).

A health monitoring start-up, using natural-language processing and machine learning, correctly predicted the spread of Covid-19 before anybody else (Niiler, 2020). Artificial intelligence (AI) was used extensively and in various forms in the context of Covid-19 (Council of Europe, 2020). AI applications were introduced to track the pandemic in real-time, to predict accurately where the virus might appear next, and to facilitate the development of an effective vaccine (Hantrais et al, 2020). AI was capable of processing vast amounts of unstructured text data to predict the number of potential new cases by area (Hantrais et al, 2020) and to forecast which types of populations would be most at risk, while also assessing, evaluating, and optimizing strategies for controlling the spread of the epidemic (Kritikos, 2020c).

But even before the pandemic, social scientists recognized that technological development and economic growth did not necessarily result in social progress (Hantrais & Lenihan, 2021).

### 1. How analyzing Google searches can support COVID-19 research

Search is often where people come to get answers on health and wellbeing, whether it’s to find a doctor or treatment center or understand a symptom better just before a doctor’s visit (Gabrilovich, 2020). In the past, researchers have used Google Search data to gauge the health impact of heatwaves, improve prediction models for influenza-like illnesses, and monitor Lyme disease incidence (Gabrilovich, 2020). Google Trends makes available a dataset of search trends for researchers to study the link between symptom-related searches and the spread of COVID-19 with the aim of a better understanding of the pandemic’s impact (see: searchingcovid19.com/, 2020). Using the dataset, researchers can develop models and create visualizations based on the popularity of symptom-related searches. Researchers could use this dataset to study if search trends can provide an earlier and more accurate indication of the reemergence of the virus in different parts of the country, but they also can be useful in studying the secondary health effects of the pandemic (Gabrilovich, 2020).

Ferguson et al (2005) show that early detection of disease activity when followed by a rapid response, can reduce the impact of both seasonal and pandemic influenza. Ginsberg et al (2009) showed the way to improve early detection and how to monitor health-seeking behavior in the form of online web search queries. They presented a method of analyzing large numbers of Google search queries in the USA to track influenza-like illness in a population. “Because the relative frequency of certain queries is highly correlated with the percentage of physician visits in which a patient presents with influenza-like symptoms, we can accurately estimate the current level of weekly influenza activity in each region of the United States, with a reporting lag of about one day “Ginsberg et al (2009).

About 90 million American adults are believed to search online for information about specific diseases or medical problems each year (Fox, 2006), making web search queries a uniquely valuable source of information about health trends. A set of Yahoo search queries containing the words “flu” or “influenza” were found to correlate with virologic and mortality surveillance data over multiple years (Polgreen et al, 2008). Ginsberg et al (2009) show that up-to-date influenza estimates may enable public health officials and health professionals to better respond to seasonal epidemics. If a region experiences an early, sharp increase in Covid-19-like-illness physician visits, it may be possible to focus additional resources on that region to identify the etiology of the outbreak, providing extra vaccine capacity or raising local media awareness as necessary (Ginsberg et al, 2009).

Google Trends (GT) uses Google to explore the searching trends of specific queries. GT may predict the outbreak of many diseases (Bousquet et al, 2017). In Germany, correlations between the patient-based, combined symptom medication score (allergy) and GT data are stronger than those with the regionally measured pollen count data (Konig, Mosqes, 2014). Search activities using GT for terms such as ‘‘allergy,’’ ‘‘allergies,’’ and ‘‘pollen’’ (Willson et al, 2015) correlate strongly with observed pollen counts. GT reflects the real-world epidemiology of symptomatic allergic rhinitis (Kang et al, 2015) and could potentially be used to monitor allergic rhinitis. Seasonality of allergic rhinitis was found using Internet searches (Marques et al, 2016) and correlated with pollen counts. Google search interests can also be used to predict the number of asthma-related emergency department visits in the area (Ram et al, 2015). In the study from 2017 Bousquet et al used GT to compare terms related to asthma, allergy, and rhinitis in 10 countries from 2004 to 2016. This study shows that seasonal asthma can be identified by GT when there is a severe asthma outbreak (Bousquet et al, 2017).

The project by Google Trends, Schema, and Axios shows how searches became more specific as infections of Covid-19 spread across the United States (Kight, 2020). This data gives an intimate look at how individuals have reacted to the uncertainty as the virus cases have risen and they find distinct regional patterns over time that reflected the spread of the virus. Coronavirus-related Google searches have surged since January, with Feb. 26 2020 the turning point at which “coronavirus” began to surpass three typical top Google searches nationally — “Facebook,” “YouTube” and “Amazon” — according to Google Trends data (Kight, 2020).

The search trends also signaled how widely people are heeding the advice from public health officials, who early on urged Americans to wash their hands, and more recently to wear masks in public. Americans across the states began searching for basic information about the virus even before it began spreading in the U.S. “What is a pandemic?” and “What are the symptoms of the coronavirus?” began rising and spreading through the U.S. in January and February (Kight, 2020). But the searches became more narrow, personal, and actionable following states’ first confirmed coronavirus case — shifting to more “how-to” queries. With the first report of a confirmed U.S. coronavirus-related death on Feb. 29, queries took on a new sense of immediacy. People sought more information about specific symptoms. They searched, “What is a dry cough” and “What is considered a fever” (searchingcovid19.com/, 2020).

### 2. Methodological explanation

The basic hypothesis of this paper is that the analytical tool Google Trends is a useful source of data for determining, estimating, and predicting an increase in the number of new active cases in the country and by regions in Croatia.

The basic methodological concept of our approach is to monitor the digital trace of language searches with the Google Trends analytical tool (trends.google.com). The GT analytics application is a trend search tool that shows the popularity of a term when searching on Google, and we can see if a trend is rising or falling. GT does not provide information on the actual number of keyword searches. Instead, it standardizes search volume on a scale of 0 to 100 over the period being examined, with higher values indicating the time when the search volume was greatest, allowing for verifiable metrics (trends.google.com).

To standardize the data, we requested the data for the period from 20. January 2020 to 20. February 2021. We then divided the keyword frequency for selected words giving us a search frequency index. We have then compared searches with official statistics to prove the significations of results (see further explanations by Wilde et al., 2020). To select the most common terms to be searched, we adhered to the WHO report (2020). Infected with Covid-19 may be asymptomatic or develop symptoms such as fever, cough, fatigue, shortness of breath, or muscle aches. A review of 55,924 laboratory-confirmed cases in China showed the following typical signs and symptoms: fever (87.9% of cases), dry cough (67.7%), fatigue (38.1%), sputum production (33.4%)., shortness of breath (18.6%), sore throat (13.9%), headache (13.6%), muscle pain or joint pain (14.8%), chills (11.4%), nausea or vomiting (5.0%), nasal congestion (4.8%), diarrhea (3.7%), hemoptysis (0.9%), and conjunctival congestion (0.8%) (WHO, 2020). Further development can lead to severe pneumonia, acute respiratory distress syndrome, sepsis, septic shock, and death.

Further, keywords were chosen by brainstorming possible words that we believed to be predictive, specific, and common enough for use in forecasting (we used a similar method in predicting migration from Croatia; see Jurić, 2020, Digital demography). After the significance screen, we selected keywords and topics.

To understand these terms, a note on the logic behind the Google Trends search algorithm is necessary (Wilde et al., 2020). Certain delimiters, such as “, -, and + allow users to change the combinations of keywords searched. A search for a single keyword will yield the search frequency index counting all searches containing that keyword, including searches that contain other words (Wilde et al., 2020).

The study we present has important limitations that we want to highlight. Although previous research in this area has shown the feasibility of using digital data for demography, at the same time we highlight the problems associated with assessments and conclusions (see: Zagheni, Weber, Gummadi, 2017; Zhang 2020). Namely, it is unquestionable that there are still significant open methodological issues and the questionable integrity of the data obtained using the sources of large data sets. Unquestionably, this model has unresolved issues related to the reproducibility of the findings and the validity of the measurements, which arise from the very characteristics of the Google Trends (GT) system used (Jurić, 2021 unpublished study). When using this tool it should be borne in mind that each of these searches was conducted for its reason and does not answer direct questions from researchers. Thus, for example, “googling” the term “coronavirus testing” is not necessarily an implication that someone is ill or experiencing symptoms. The search queries are not exclusively submitted by users who are experiencing Covid 19 symptoms, and the correlations we observe are only meaningful across large populations. However, testing of a similar model used in the U.S. for the onset of influenza showed a high correlation between influenza symptom searches and physical reports of influenza cases (Ginsberg et al, 2009).

The fact is also a problem that GT does not provide data on which population was sampled or how it was structured. Despite strong correlations, this system remains susceptible to false alerts. An unusual event, such as a drug recall for a popular cold or flu remedy, could cause such a false alert. (Ginsberg et al, 2009). Although these open-ended issues pose serious challenges for making clear estimates, statistics offer a range of tools available to deal with imperfect data as well as to develop controls that take data quality into account (see: “R”; see Zagheni, Weber, Gummadi, 2017).

We emphasize that this system is not designed to be a replacement for traditional surveillance networks or supplant the need for laboratory-based diagnoses and surveillance.

None of the queries in this project’s database can be associated with a particular individual. This project’s database retains no information about the identity, IP address, or specific physical location of any user.

### 3. Occurrence and spread of coronavirus in Croatia

The first case of SARS-CoV-2 virus infection in Croatia was confirmed on February 25, 2020. (koronavirus.hr). In mid-February 2021, there were 237,725 patients in the Republic of Croatia, while a total of 1,268,323 people was tested for coronavirus, and 11,694 people were in self-isolation (koronavirus.hr). On March 11, 2020, a Decision was made in Croatia to declare an epidemic of the COVID-19 disease caused by the SARS-CoV-2 virus. This decision declared an epidemic in the entire Republic of Croatia on 11 March 2020 (zdravlje.hr, 2020).

On the same day, the World Health Organization declared a previous epidemic a pandemic (WHO, 2020). On March 22, 2020, Croatia was hit by an earthquake measuring 5.5 on the Richter scale (pmf.unizg.hr, 2020), which is the strongest earthquake in Zagreb after the 1880 earthquake. It is assumed that the consequences caused by the earthquake contributed to the behaviors that contributed to the increase in the number of newly infected in the coming period.

On April 2, more than 1,000 infections were recorded in Croatia. In the next three weeks, the number has already doubled (koronavirus.hr). Since May, the number of newly infected has been declining rapidly. On May 10, the Civil Protection Headquarters announced that passes were being abolished in Croatia. (civilna-zastita.gov.hr, 2020). In the period from May 30 to June 3, no new cases were recorded. On July 10, 116 new cases were recorded, which was the highest daily number of newly infected since the beginning of the epidemic in Croatia (koronavirus.hr). Notice on keeping a safe distance of at least two meters (HZJZ.hr, 2020). On August 8, 77 new cases were recorded, and at the end of August, the then-record number of 358 new cases was recorded (HZJZ.hr, 2020). Due to the increase in the number of patients per 100,000 inhabitants, Croatia is on the red list of a total of 13 EU countries.

**Figure 1.**
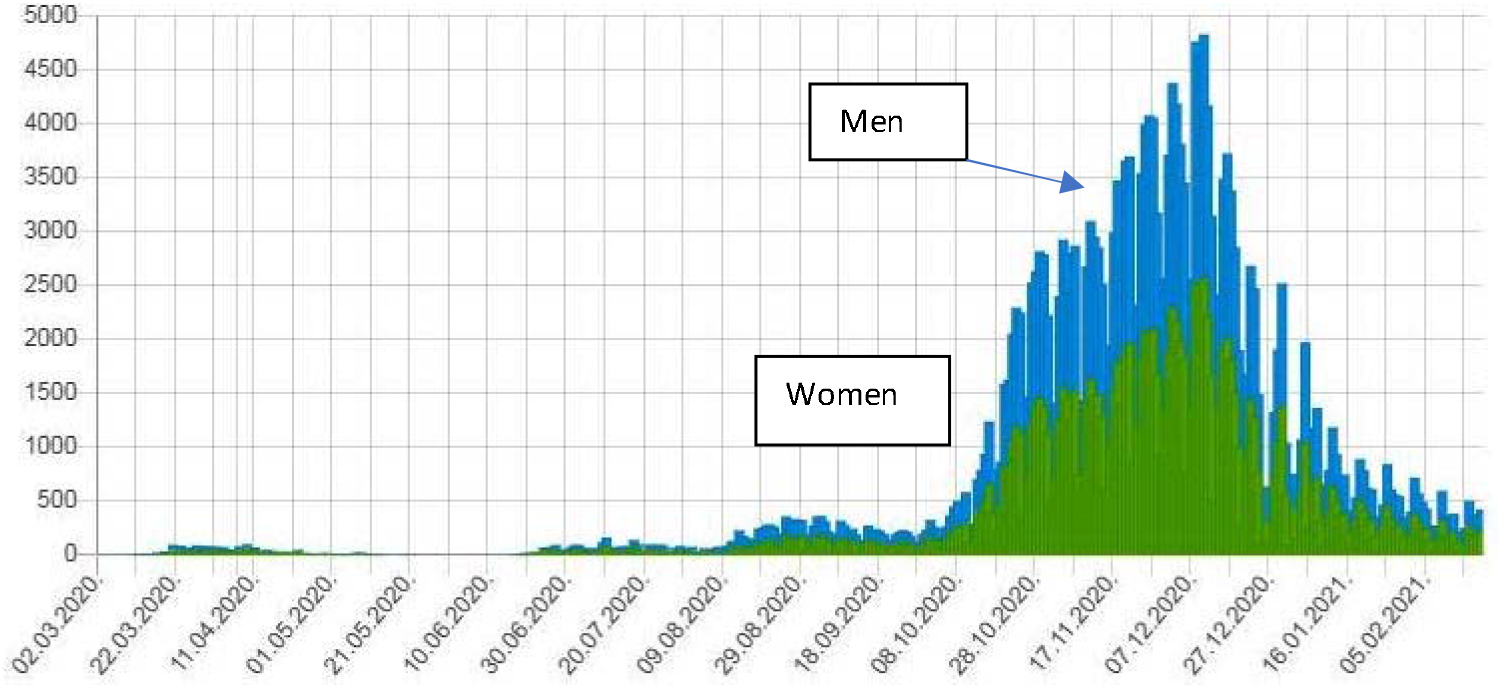
Number of total patients (active cases) in Croatia, March 2020 – February 2021. Izvor: HZJZ, 2021

**Figure 2.**
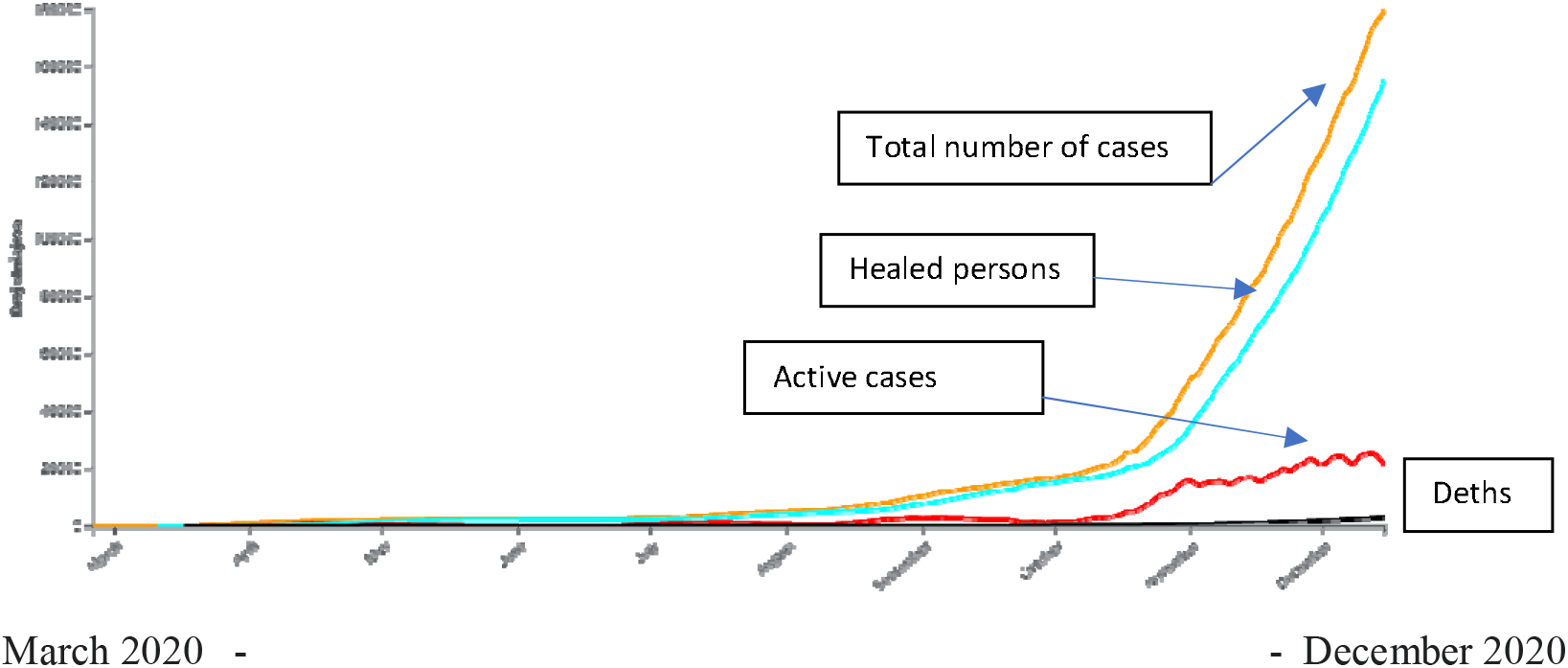
Number of total patients (active cases) in Croatia, March 2020 – February 2021. At the beginning of September, a record number of 369 new cases was recorded, and in mid-October, 793 new cases. During October, the number doubles, and by the end of October it triples (koronavirus.hr, 2020). Source: koronavirus.hr, 2020)

Due to the dramatic increase, the National Civil Protection Headquarters has announced new strict measures to combat the epidemic (civilna-zastita.gov.hr, 2020). On Nov. 6, a new record of 2,890 new cases was recorded in 24 hours. The number of active cases was 15,567 (koronavirus.hr).

**Figure 3.**
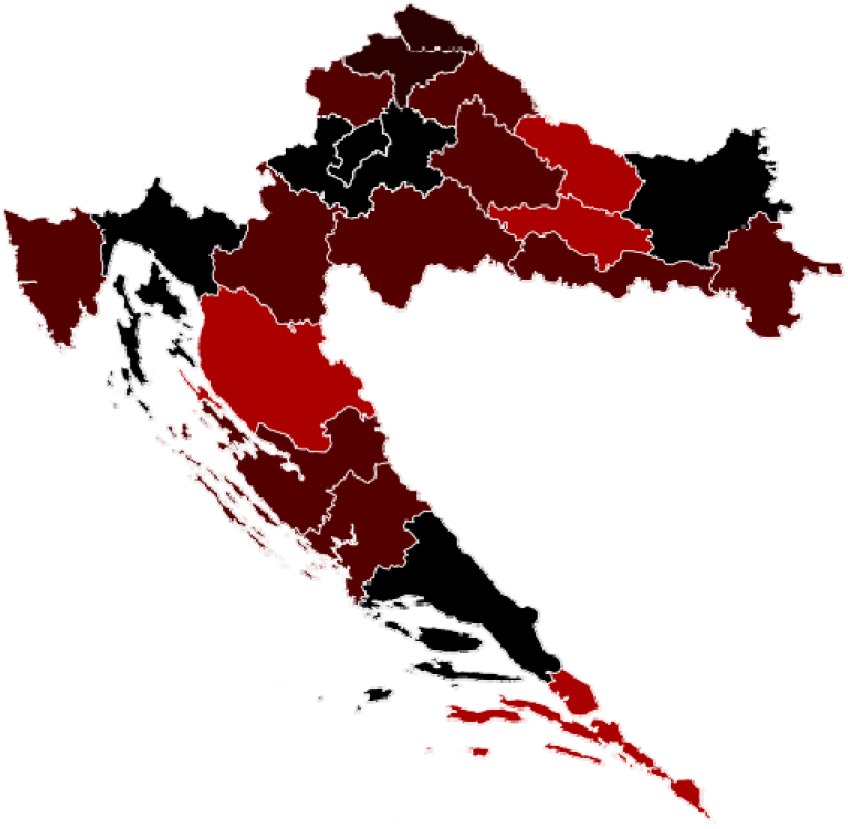
Reported cases in Croatia by region, March 2020 – February 2021. Legend: Darker shades indicate a higher number of cases Source: koronavirus.hr, 2020)

Since mid-December, the situation has improved significantly and there are fewer active and new cases every day. In the next part of the year, the situation will change, and by January 2021, with just over 1,000 registered cases per million inhabitants related to the coronavirus pandemic (a total of 4,403 registered cases by the first week of 2021), Croatia will record data similar to most other countries in EU (koronavirus.hr, 2021). The situation remains the same in the following period, so on 12 February 2021, the Government of the Republic of Croatia adopted a Decision amending the Decision on Necessary Epidemiological Measures (OG 2021), which mitigates anti-epidemic measures.

By the end of September, according to estimates by the Croatian Institute of Public Health, approximately 10 percent of the population, ie. about 400,000 people were infected with coronavirus and developed antibodies (HZJZ.hr, 2021). The so-called “herd immunity”, ie. a situation where the infection can no longer spread strongly in the population due to a sufficient number of individuals with developed immunity is expected for the beginning of summer 2021.

In the first 400,000 people infected with coronavirus, a total of 257 patients died (koronavirus.hr, 2021). According to the University of Oxford, at the beginning of 2020, Croatia was among the countries with the strictest restrictions and measures to reduce infection with the new coronavirus (VL, 2021).

At the end of 2020 and the beginning of 2021 there is a strong fall in the number of deaths due to pandemics and newly registered cases of infection, which some scientists attribute to the introduction of tougher anti-epidemiological measures in late 2020, and others to the fact that a significant percentage of the population was infected with coronavirus and developed antibodies (T-portal.hr, 2021).

### 4. Results

The Google search Index cannot estimate the exact number of searches, so with the help of this tool the exact number of new cases cannot be estimated, but the increase of the trend can be noticed very precisely, which can serve as an indicator of new cases in the whole country and individual regions. Its main advantage compared to official indicators is that it detects the phenomenon as quickly as possible and thus can serve as an early alarm.

With the outbreak of pandemic in Croatia and lockdown in March 2020 Croatian citizens are beginning to googling intensively for terms related to Covid-19. They searched, “What is a dry cough”, “What is considered a fever”, “coronavirus” etc. We checked those Covid-19 related queries in Croatia and noticed the following.

**Figure 4.**
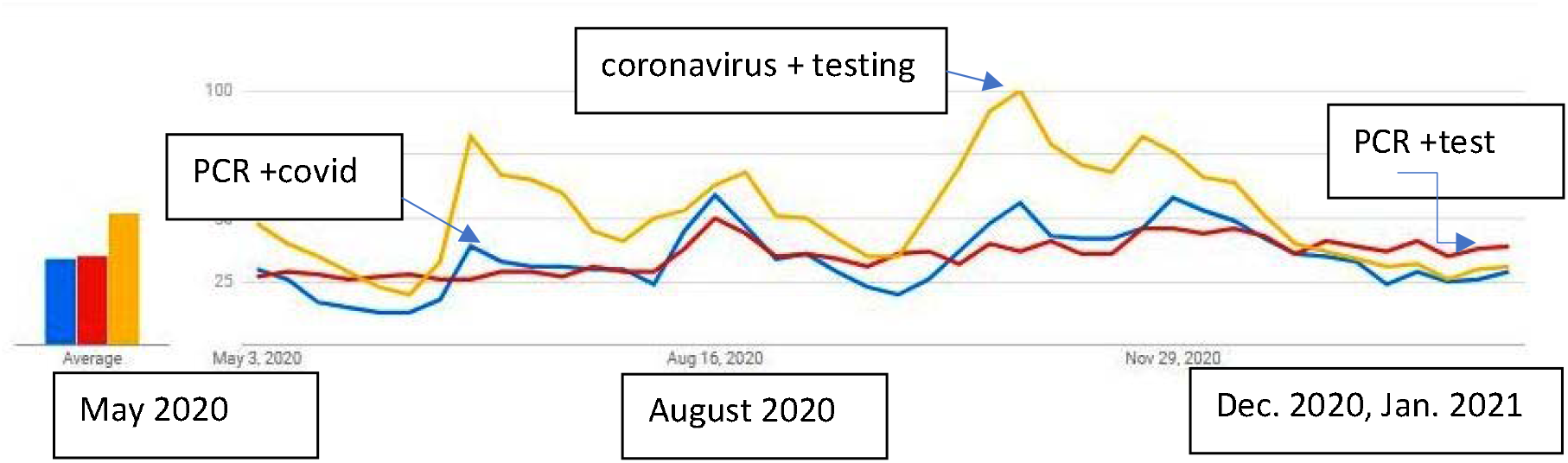
Queries reported by GT concerning queries “PCR +Covid”, “PCR + test” and “coronavirus + testing” from May 2020 to February 2021 in Croatia. The Increase in Google search queries “PCR +Covid”, “PCR + test” and “coronavirus + testing” is correlated with the increase in the number of new cases. The decrease in Google search is correlated with the decrease in the number of new cases.

**Figure 5.**
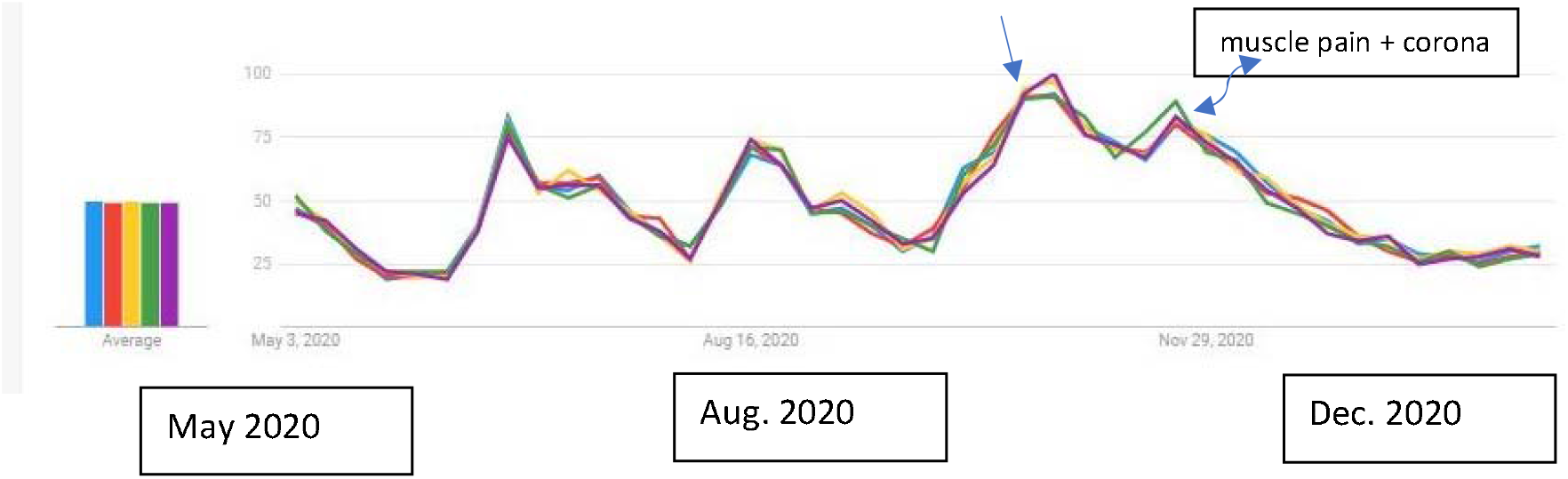
Queries reported by GT concerning symptoms “cough + corona”; “pneumonia + corona”; “dry cough + corona”; “runny nose + coronavirus”; “muscle pain + corona” from May 2020 to February 2021 in Croatia. The Increase in Google search queries “cough + corona”; “pneumonia + corona”; “dry cough + corona”; “runny nose + coronavirus”; “muscle pain + corona” is correlated with the increase in the number of new cases. The decrease in Google search is correlated with the decrease in the number of new cases.

**Figure 6.**
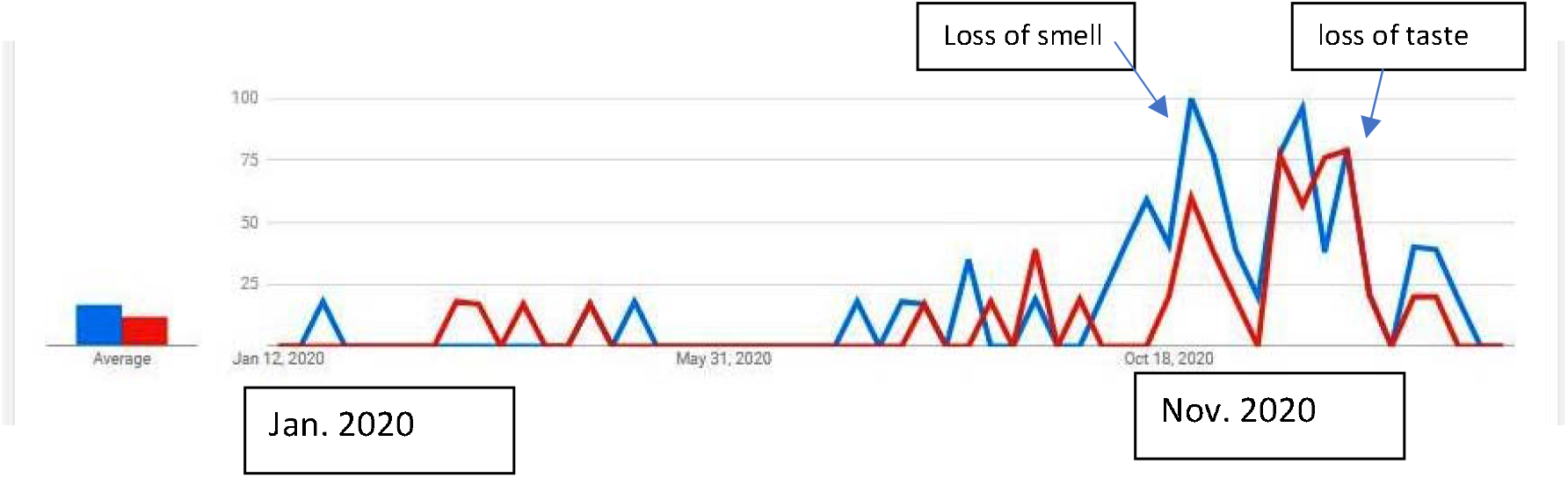
Queries reported by GT concerning “loss of smell” and “loss of taste” from January 2020 to February 2021 in Croatia. The Increase in Google search queries “loss of smell” and “loss of taste” is correlated with the increase in the number of new cases. The decrease in Google search is correlated with the decrease in the number of new cases.

**Figure 7.**
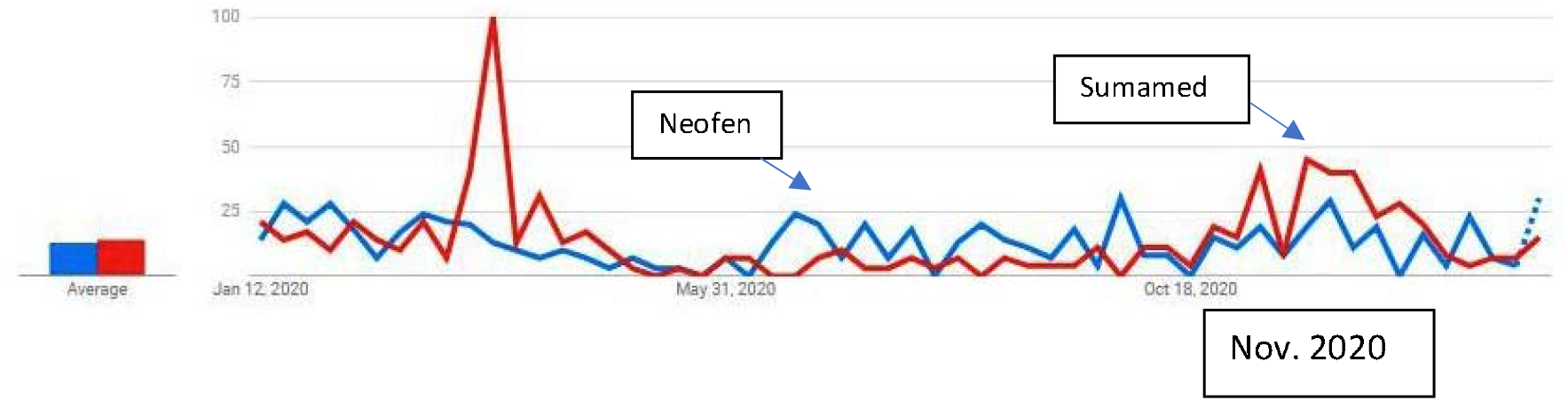
Queries reported by GT concerning “Neofen” (medication) and “Sumamed” (Cro. most popular antibiotic) from January 2020 to February 2021 in Croatia. We studied also queries “Neofen” (medication) and “Sumamed” (Cro. most popular antibiotic). It can be seen that the demand for these drugs increased especially at the time of the pandemic outbreak and in the fall of 2020 when the largest number of new cases was recorded.

Google search interests can also be used to predict the number of Covid-19-related emergency department visits in the area.

**Figure 8.**
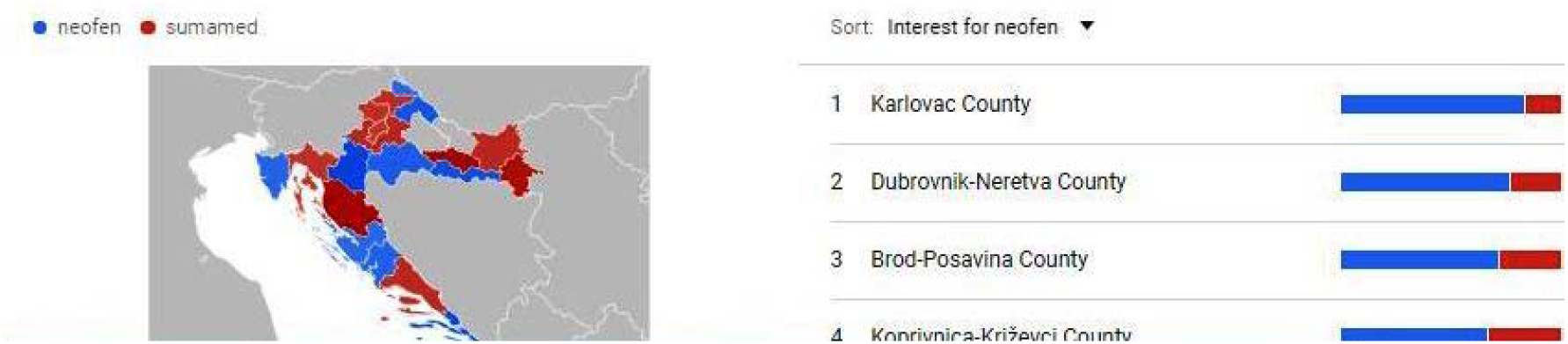
Correlation by regions. The search for these drugs correlates with a sharp increase in the number of new cases in November 2020 in the northern Croatian regions (Novi list.hr, 2020).

In further proceedings to standardize the data, we requested the data for the period from January 2020 to 20. February 2021 and divided the keyword frequency for each word (see table 1) giving us a search frequency index. Then we have compared searches with official statistics to prove the significations of results (see HZJZ.hr). We especially focused on the so-called second wave of infection spread when cases began to grow exponentially.

**Table 1:** Keyword and topic selection criteria.

| Symptoms | What is/How to? | Terms | Activities |
| --- | --- | --- | --- |
| Asthma | What is coronavirus? | Influenza Complication | Coronavirus testing |
| Anosmia | "What is Zoom?" | Cold/Flu Remedy | PCR test |
| Common cold | How to make a face mask? | General Influenza Symptoms | Application for a minimum wage |
| Cough | How to make hand sanitizer? | Term for Influenza |  |
| Depression |  | Symptoms of an Influenza Complication |  |
| Fatigue |  | Antibiotic Medication |  |
| Fever |  | Remedies for corona |  |
| Headache |  | General Influenza |  |
| Nausea |  | Antiviral Medication |  |
| Shortness of breath |  |  |  |

**Figure 9.**
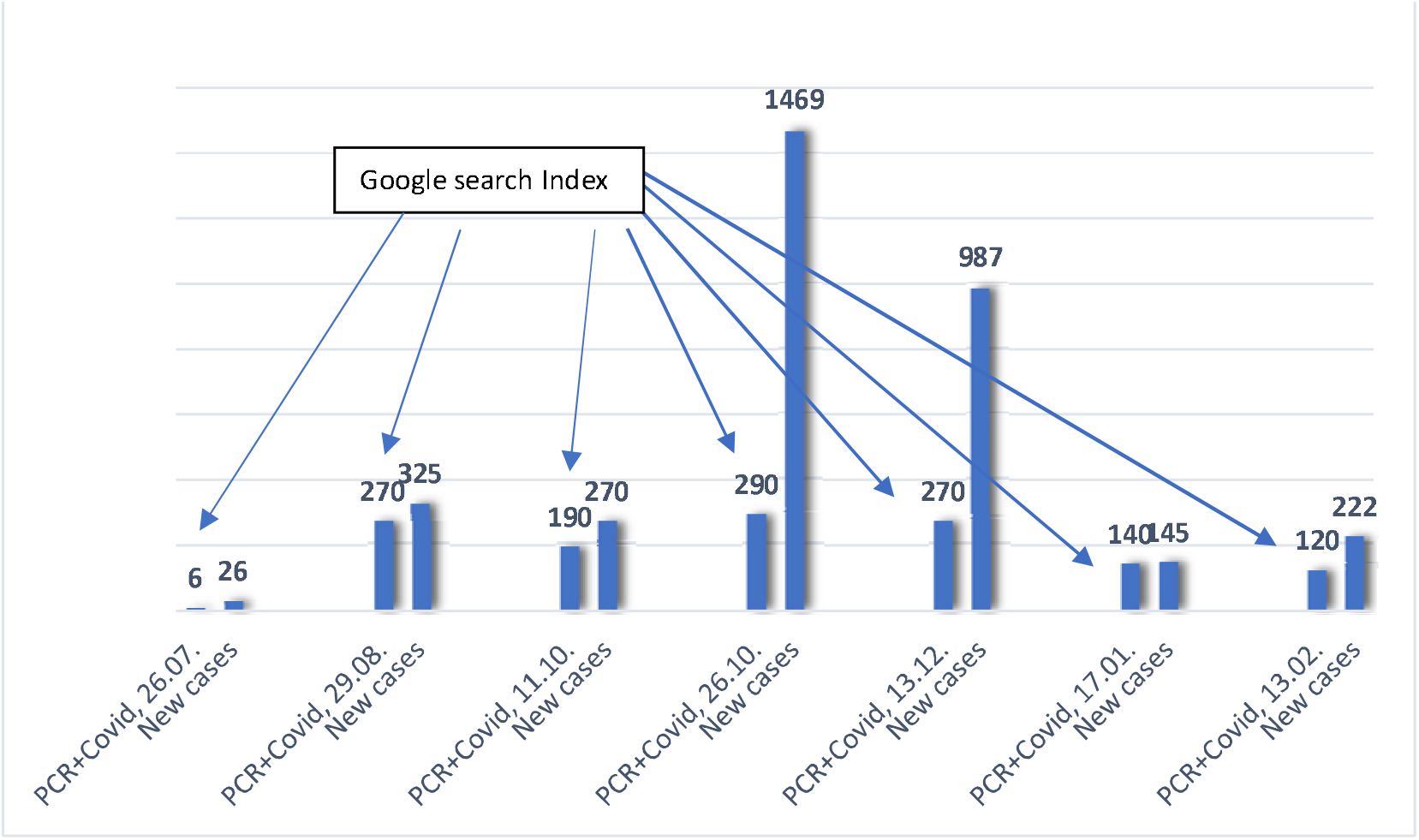
Correlation between Google search Index for query “PCR +Covid” and the official number of reported new cases of Covid-19 patients in Croatia by selected dates 2020 and 2021. The graph shows that the increase in Google search is clearly correlated with the increase in the number of new cases and that the decrease in Google search is correlated with the decrease in the number of new cases.

**Figure 10.**
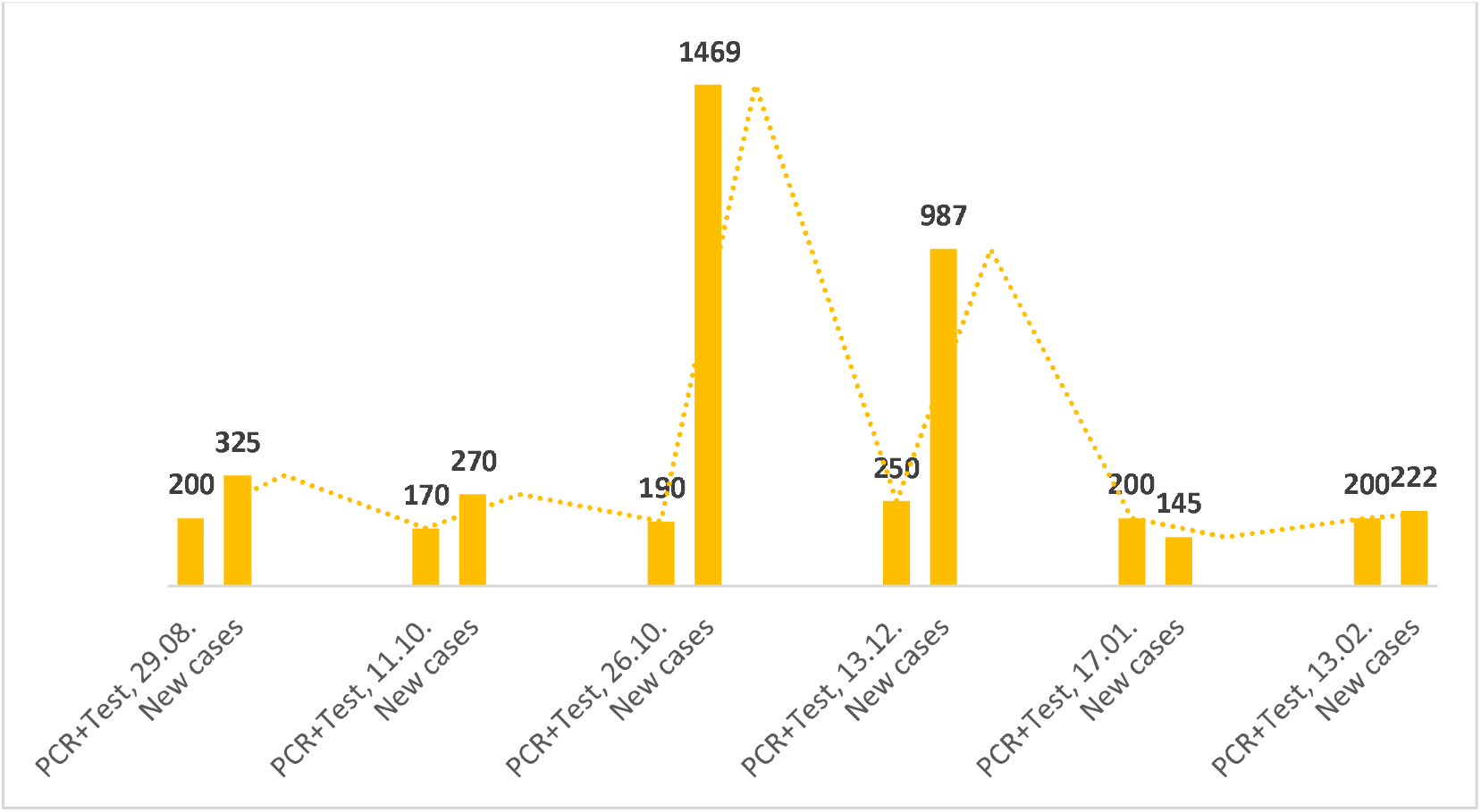
Correlation between Google search Index for query “PCR +Test” and the official number of reported new cases of Covid-19 patients in Croatia by selected dates 2020 and 2021.

**Figure 11.**
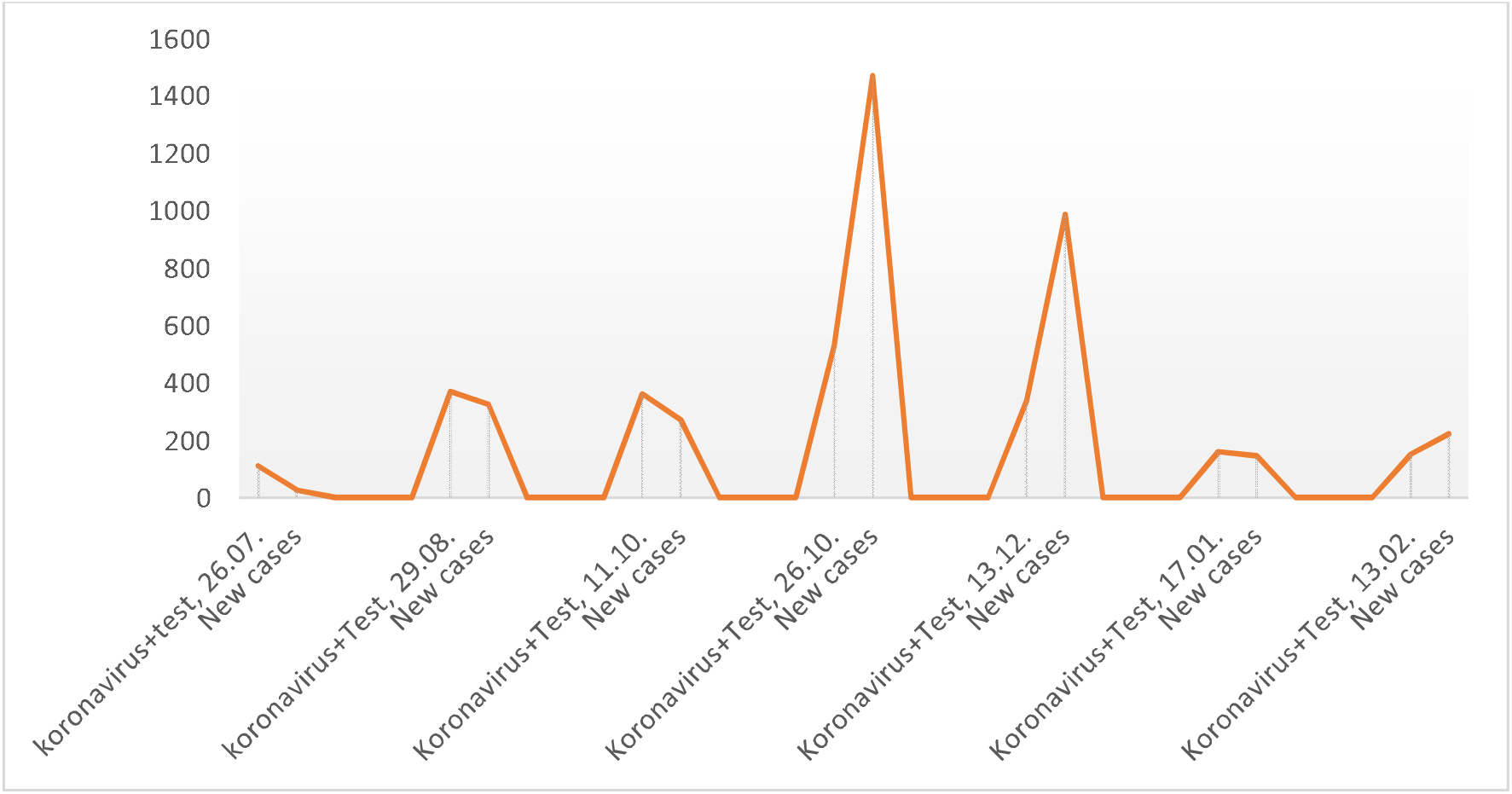
Correlation between Google search Index for query “coronavirust+test” and the official number of reported new cases of Covid-19 patients in Croatia by selected dates 2020 and 2021. All search activities using Google for significant terms ‘‘PCR +covid”, “PCR + test”, “coronavirus + test” correlate strongly with observed official cases of the disease (see HZJZ.hr).

In the continuation of the work we tested the model for most common symptoms cough, pneumonia and muscle pain and compared the data with official data (see HZJZ.hr).

**Figure 12.**
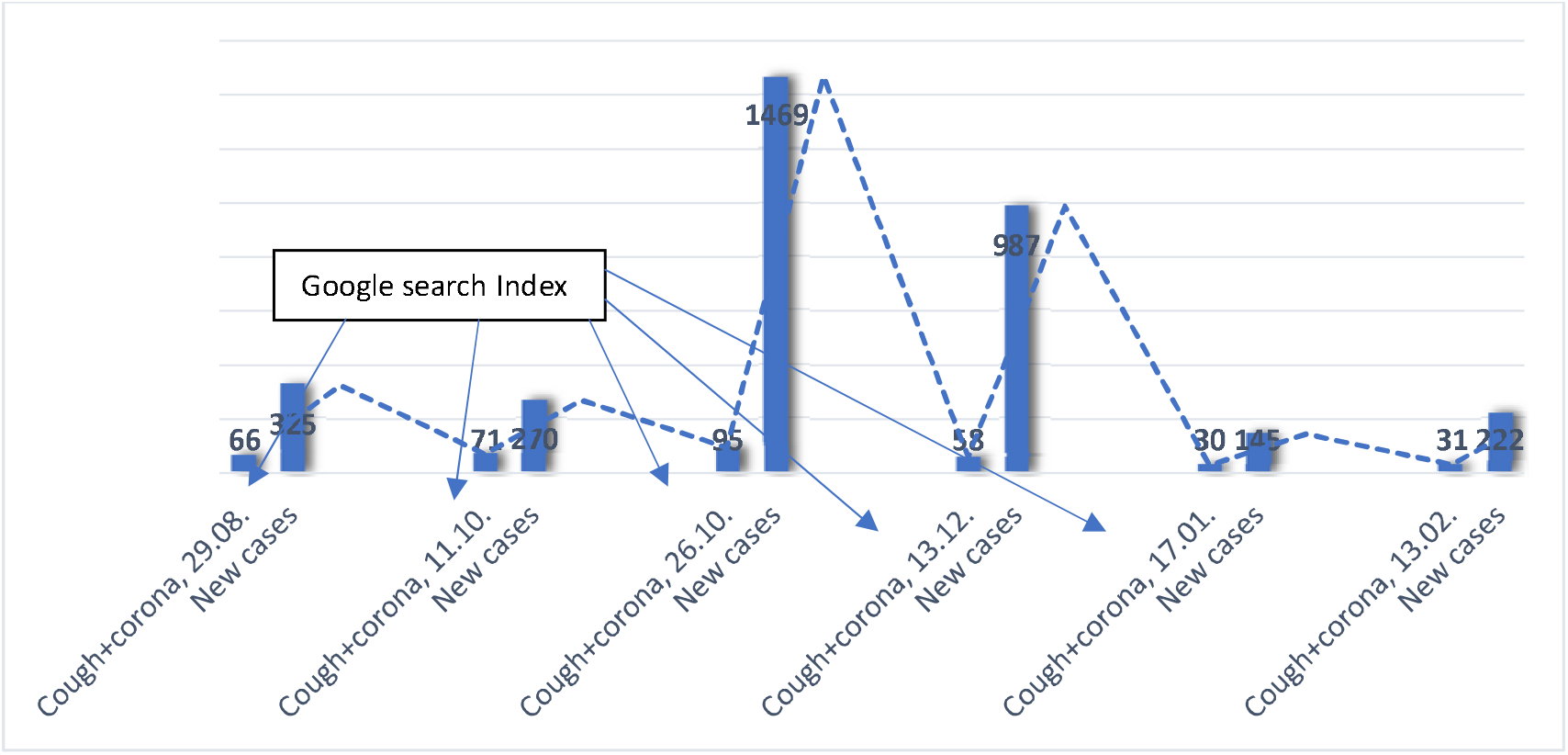
Correlation between Google search Index for query “cough +coronavirus” and official number of reported new cases of Covid-19 patients in Croatia by selected dates 2020 and 2021.

**Figure 13.**
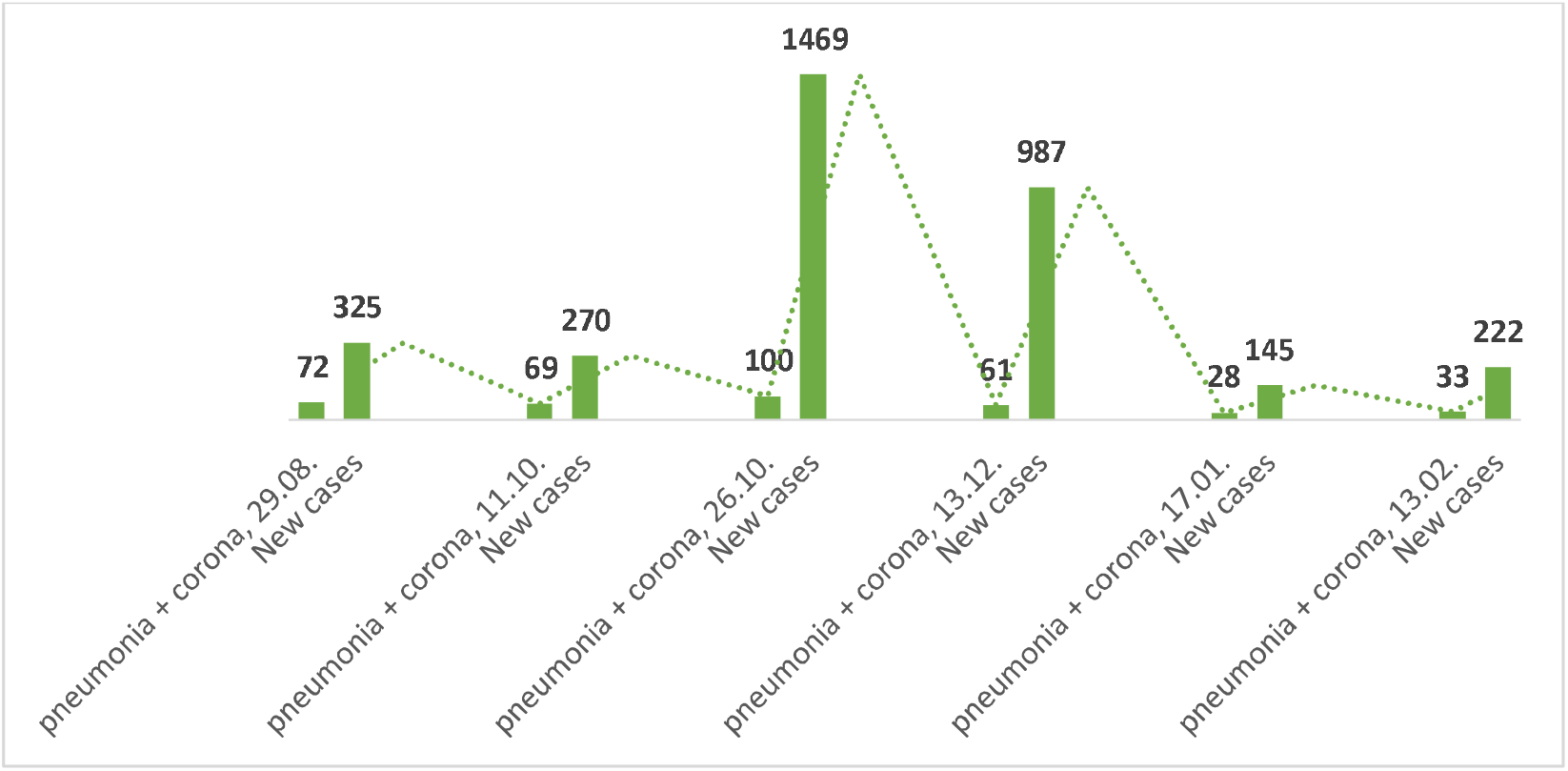
Correlation between Google search Index for query “pneumonia +corona” and the official number of reported new cases of Covid-19 patients in Croatia by selected dates 2020 and 2021.

**Figure 14.**
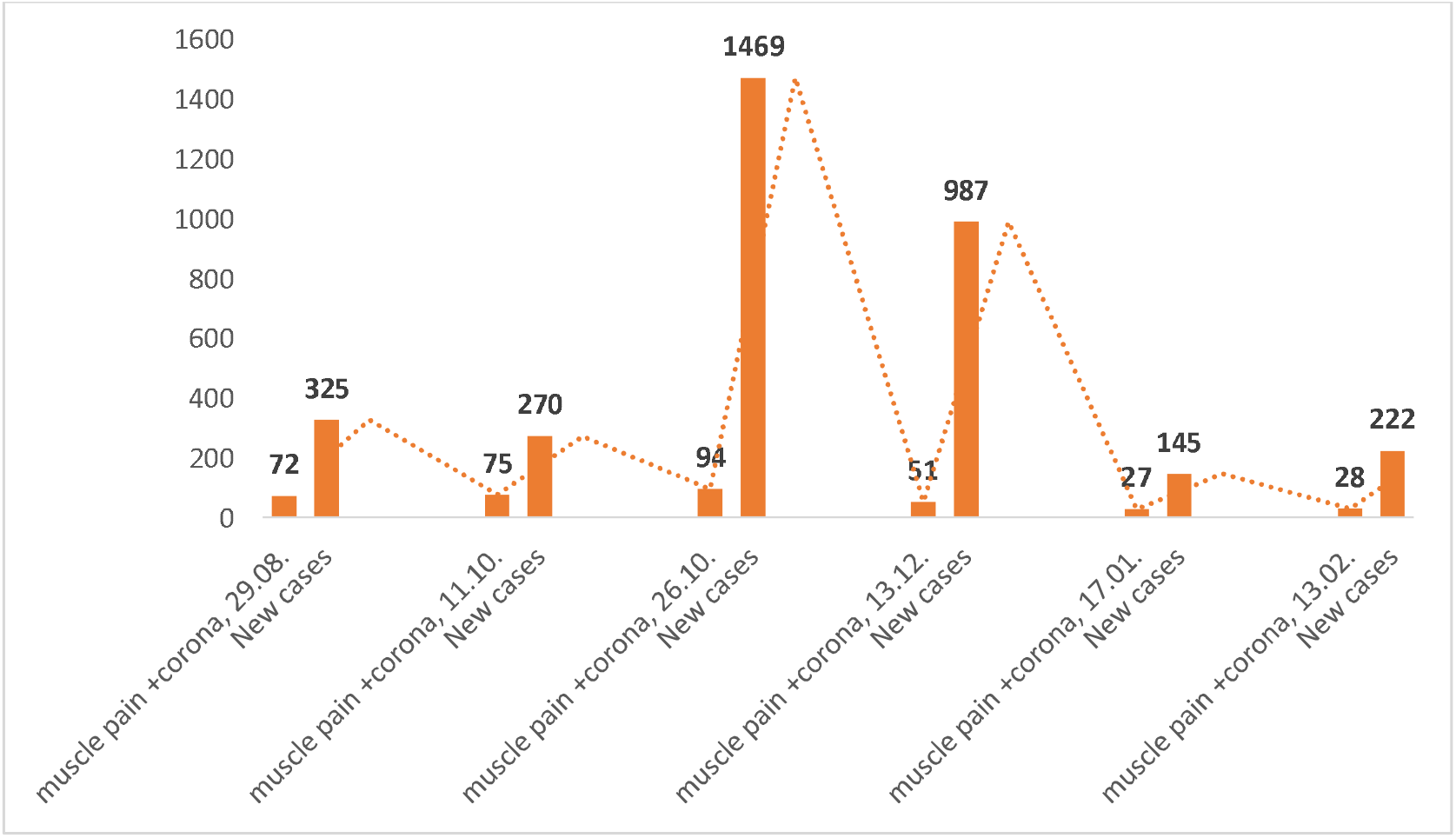
Correlation between Google search Index for query “muscle pain +corona” and the official number of reported new cases of Covid-19 patients in Croatia by selected dates 2020 and 2021. The search activities using Google for symptoms “cough + corona”, “pneumonia + corona”; “muscle pain + corona” correlate strongly with official data of new cases.

All graphs show that the increase in Google search is correlated with the increase in the number of new cases and that the decrease in Google search is correlated with the decrease in the number of new cases.

## Conclusion

Google Trends can provide one of the most timely, broad-reaching Covid-19 monitoring systems available today. As with other syndromic surveillance systems, the data are most useful as a means to spur further investigation and collection of direct measures of disease activity.

This tested method shows that the increase in Google search is correlated with the increase in the number of new cases and that the decrease in Google search is correlated with the decrease in the number of new cases. Researchers could use this dataset to study if search trends can provide an earlier and more accurate indication of the reemergence of the virus in different parts of the country. This method may enable public health officials and health professionals to better respond to seasonal epidemics. If a region experiences an early, sharp increase in Covid-19-like-illness physician visits, it may be possible to focus additional resources on that region to identify the etiology of the outbreak, providing extra vaccine capacity or raising local media awareness as necessary.

## Data Availability

In our work, we use only anonymous, aggregate data. All data are collected following the applicable GDPR and ethical principles of personal data handling.

## Funding

This research received no external funding.

